# Comparative Analysis of the Chemotherapy-related Cognitive Impairments in Patients with Breast Cancer: a Community-based Research

**DOI:** 10.1101/2021.08.23.21262481

**Authors:** Maryam Owrangi, Mohammad Javad Gholamzadeh, Maryam Vasaghi Gharamaleki, Seyedeh Zahra Mousavi, Ali-Mohammad Kamali, Mehdi Dehghani, Prasun Chakrabarti, Mohammad Nami

## Abstract

**Purpose:** With increasing breast cancer (BC) survival rates, the survivors’ quality of life (QoL) has become an important issue. Chemotherapy-induced cognitive impairment, known as “chemobrain” has been addressed recently. Therefore, cognitive function as one of the determinants of QoL should be considered while prescribing chemotherapeutics. In this study, we aimed to evaluate the effects of two common chemotherapy regimens on BC survivors’ cognition.

**Methods:** The participants comprised 35 BC patients who underwent two common chemotherapy regimens, AC-T and TAC, and 24 matched healthy volunteers. The participants were assessed regarding anxiety, depression, general health status, and cognitive function including aspects of concentration, verbal ability, reasoning, memory, and visuospatial skill through Addenbrooke’s Cognitive Examination (ACE-P) and Cambridge Brain Science (CBS) tests.

**Results:** Regarding depression and anxiety, there were no significant differences between the three groups. However, BC patients significantly complained of chronic fatigue compared to healthy volunteers (P-value = 0.027). Besides, ACE-P revealed the language domain to be affected in the AC-T group in comparison with the TAC-treated cases (P-value = 0.036). Moreover, the patients receiving the AC-T regimen had worse performance in visuospatial working memory and attention domains compared to the TAC group considering CBS tests (P-value = 0.031 and 0.008, respectively).

**Conclusion:** The results represent the AC-T regimen to be more toxic than the TAC in domains of language, concentration, and visuospatial working memory.

**Implications for cancer survivors:** The AC-T regimen should be prescribed with caution in BC patients suffering from baseline cognitive impairments to improve post-chemotherapy QoL.

**Graphical Abstract:** 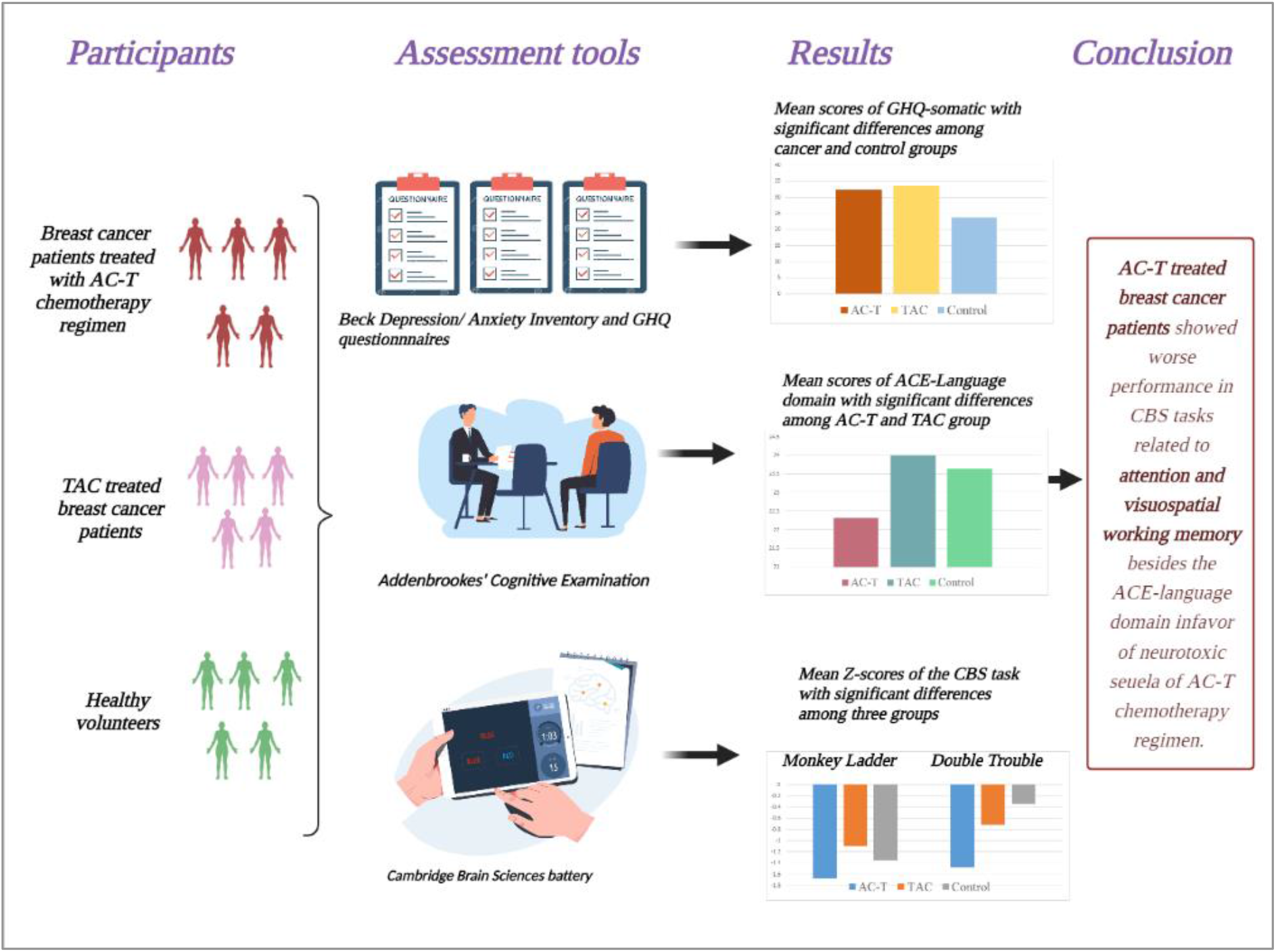

## 1. Introduction

Breast cancer (BC) is the most common cancer and the leading cause of cancer deaths among women [1]. In recent years, with the early detection of the patients and the development of treatment methods, the number of survivors of this disease has increased in many countries [2, 3]. With the increasing number of disease-free survivors, the concept of Quality of life (QoL) of BC survivors becomes important. Although BC survivors reported lower QoL than the general population immediately after diagnosis and treatment, their QoL improved over time and even some researches showed they have the same QoL as normal age-matched controls but they still suffer from symptoms such as cognitive problems, fatigue, insomnia, sexual issues, etc. [4-8].

As mentioned above, the cognitive deficit in BC survivors may be presented for many years and affect their QoL. This chemotherapy-induced cognitive impairment is experienced by many BC patients after treatment and is known as “chemofog” or “chemobrain” in literature. The prevalence of chemobrain has been reported differently in some articles. While self-reported cognitive dysfunction prevalence is up to 70% [9-11], there is no association between perceived and objective cognitive impairment in cancer survivors, and even some studies showed there is no objectively detected cognitive decline [9, 11, 12]. Hutchinson et al. suggested that these discrepancies could be explained by variation in cognitive assessments and definition of impairment or the hypothesis that ‘subjective cognitive impairment may be an indicator of psychological distress rather than chemotherapy-induced cognitive impairment’ [13]. Another possible explanation is that chemotherapy causes mild cognitive impairment which is not detected by neuropsychological tests but affects the patients’ daily life [14].

Studies have revealed that chemobrain acts on different cognitive domains including visuospatial skill, attention, delayed memory, processing speed, executive functions, and concentration with various severity [9, 15-17]. For instance, Jim et al. indicated that chemotherapy causes mild deficit only in verbal and visuospatial domains [18]. The other study by Henderson et al. reported impairment in memory, language, and processing speed [19]. Li and his colleagues in their review illustrated that attention, processing speed, verbal memory, and executive control are the affected domains [20].

To date, the exact mechanism of chemobrain is not well understood. However, it is established that this phenomenon is multifactorial and factors like the chemotherapy regimen, patients’ age, ethnicity, socio-economic status, stage of disease, menopausal status, and psychological symptoms are possible predisposing factors [21-23]. It is demonstrated that psychological features are more important than demographic and medical features in self-reported cognitive decline [23-25]. Besides, although different chemotherapy regimens are currently prescribed, limited studies have evaluated the magnitude of cognitive impairment associated with each regimen.

Therefore, since neurotoxic consequences of each commonly prescribed chemotherapy regimen are not fully understood, in our study, we aim to compare both perceived and objective cognitive impairments of two common chemotherapy regimens and an age-matched control group besides evaluating the patients’ anxiety, depression, fatigue, socio-economic status, etc. as confounding factors that can affect cognitive impairment.

## 2. Materials and Methods

### 2.1 Participants

The study was carried out among BC patients who underwent adjuvant chemotherapy (n=35) and healthy women (n=24) between 2018 and 2020. Patients were selected from 30-65 year-old women referring to Motahari Polyclinic, the affiliated medical center of Shiraz University of Medical Sciences, Iran. Inclusion criteria consist of: A) diagnosis of BC confirmed by tru-cut biopsy, B) the last cancer treatment session (chemotherapy or radiotherapy) has been at least six months before this study, and C) having a diploma or above degrees. Exclusion criteria were: A) any prior history of diagnosed neuropsychological disorders (including major depressive disorder, bipolar disorder, schizophrenia, epilepsy, brain tumor, etc.), B) stage IV of cancer, and C) presence of brain metastasis.

Patients were divided into two groups according to their chemotherapy regimen: A) AC-T: Doxorubicin (Adriamycin®) and Cyclophosphamide (Cytoxan®) every three weeks for four cycles followed by Paclitaxel (Taxol®) for four other cycles, B) TAC: Docetaxel (Taxotere®), Doxorubicin (Adriamycin®), and Cyclophosphamide (Cytoxan®) every three weeks for six cycles.

The control group was selected from 30-65 year-old women who were referred for mammographic evaluation of benign breast nodules. They didn’t have any prior history of malignancy or neuropsychological disorders. They also were group-matched with patient groups according to age, education, and socio-economic status.

Before enrolling in the study, all participants were informed of the purpose and process of the study and signed informed consent.

### 2.2 Study Design

After selecting participants, they were invited to undergo several tests in the Neuroscience Laboratory, Department of Neuroscience, Shiraz University of Medical Sciences. They were asked to fill four questionnaires (demographic, General Health Questionnaire (GHQ), Beck Depression Inventory (BDI), and Beck Anxiety Inventory (BAI)). After that, Addenbrooke’s Cognitive Examination (ACE) was performed. Finally, they were asked to complete the Cambridge Brain Sciences (CBS) cognitive battery.

### 2.3 Assessment Tools

#### 2.3.1 Demographic questionnaire

The demographic questionnaire was designed by the authors and includes the following information: age, marital status, education, occupation, salary, past medical history and neuropsychiatric disorders of the participants and their family, medication history, substance and alcohol consumption, and smoking habits. Participants were asked about their menopausal status (in the patient groups, before and after treatment). A detailed history of the breast cancer including date of diagnosis, type of treatment, and post-treatment complications was acquired from the patient groups. Finally, they filled a 0-10 Visual Analogue Scale (VAS) for evaluation of cognitive disabilities such as any impairments in memory, attention, learning, speech, vision, judgment, problem solving and decision making, depressive and anxiety symptoms, and fatigue in such a way that a 0 score represents no problem in that cognitive domain and a 10 score is indicative of a severely disabling problem.

#### 2.3.2 General Health Questionnaire-28 (GHQ-28)

We used a verified Persian version of GHQ-28. This includes 28 questions and aims to assess mental health and consists of four domains of somatic symptoms, anxiety and insomnia, social dysfunction, and severe depression [26].

#### 2.3.3 Beck Depression Inventory (BDI) and Beck Anxiety Inventory (BAI)

All participants were asked to fill a verified Persian version of BDI and BAI. BDI is a 21-question Likert scale for the assessment of depression [27]. Similarly, BAI includes 21 questions for assessing anxiety symptoms [28].

#### 2.3.4 Addenbrooke’s Cognitive Examination - Persian version (ACE-P)

ACE is an interview-based examination designed for screening and diagnosis of dementia and other cognitive disorders. This is a more extensive diagnostic test compared to the Mini Mental State Examination (MMSE). ACE-P is a Persian version of the ACE-Revised version. The validity and reliability of ACE-P were verified for the Iranian population. Completing this test took about 30 to 40 minutes for each participant. The total score for this test is 100 points and contains five subtests, representing five cognitive domains; attention and orientation (18 points), memory (26 points), fluency (14 points), language (26 points), and visuospatial skill (16 points) [29].

#### 2.3.5 Cambridge Brain Sciences (CBS) Battery

CBS battery is a web-based platform (https://www.cambridgebrainsciences.com/) for the assessment of cognitive function which uses a battery of computerized tasks. These tasks were designed to assess visuospatial working memory (Monkey Ladder), spatial short-term memory (Spatial Span), working memory (Token Search), episodic memory (Paired Associates), mental rotation (Rotations), visuospatial processing (Polygons), deductive reasoning (Odd One Out), verbal short-term memory (Digit Span), attention (Feature Match), and response prohibition (Double Trouble) [30]. Planning (Spatial Planning) and verbal reasoning (Grammatical Reasoning) tests were excluded from our study due to the difficulty of our participants to perform these tasks. We showed the participants how each task is performed. Then they were asked to complete the tasks by themselves. Final scores of participants for each test were converted to Z-score values through default CBS metrics to compare.

### 2.4 Statistical Analysis

Data were analyzed using IBM SPSS software version 26. First, the normality of variables was assessed. For variables with normal distribution paired-sample T test, independent-sample T test, and ANOVA were used. For those without normal distribution, non-parametric equivalent tests such as two independent samples and K independent samples tests were used.

## 3. Results

### 3.1 Demographic information

In total, 59 participants were recruited in our study of whom 24 participants were healthy volunteer women as the control group and 35 responders were BC survivors who had received their last chemotherapy sessions at least 6 months prior to the enrollment. 16 participants of these 35 BC patients had been treated with AC-T regimen and the rest had received the TAC chemotherapy regimen. All of these patients had undergone mastectomy and radiotherapy as parts of their treatment protocol. Besides, all the three groups were matched regarding age, educational level, and socioeconomic status. It is noteworthy that none of the participants had a positive history of brain surgery, stroke, epilepsy, and other neurological disorders. The mean ages of the participants were 48, 44.6, and 45 years for the AC-T, TAC, and control groups respectively. The table below illustrates some of the main demographic characteristics of the participants (Table 1).

**Table 1:**
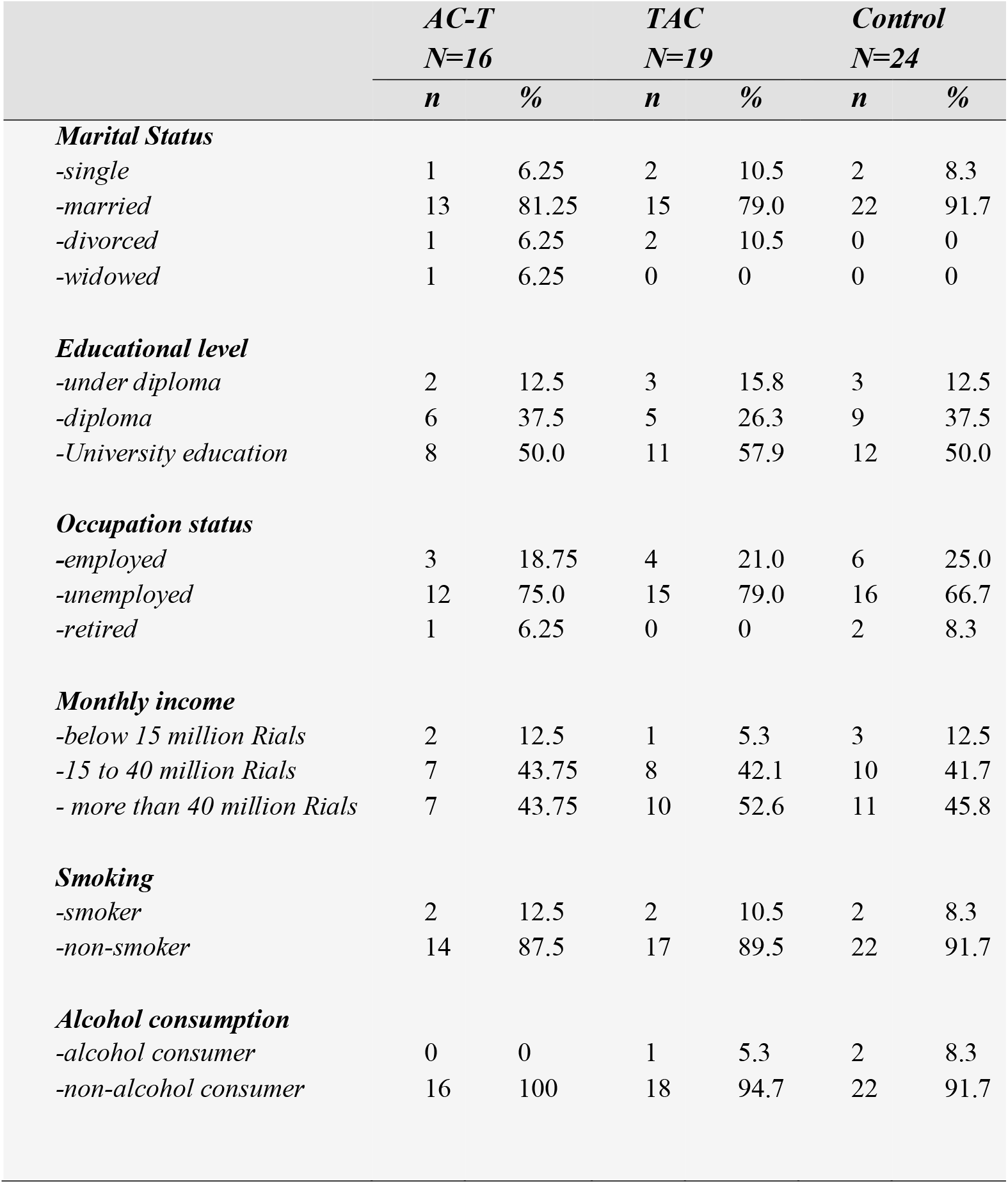
summary of demographic information of the participants.

### 3.2 Subjective cognitive impairment, psychological problem, and fatigue

With respect to assessing cognitive impairment subjectively, the participants were asked to score the severity of their cognitive complaints including the memory, attention, learning, speech, vision, and judgment disorders with a 0-10 VAS. Moreover, self-reported depression, anxiety, fatigue, and current fatigue were assessed. It was demonstrated that only subjective fatigue complaints had shown significant differences between cancer and control groups (P-value = 0.027). The mean self-reported fatigue score of the cancer group -AC-T and TAC together-was 4.06 in comparison with 2.04 of the control group. The degree of cognitive impairments in domains including memory, attention, learning, judgment, etc. did not show significant differences among the three groups subjectively.

### 3.3 GHQ, BAI, and BDI

As mentioned above, assessment of depression, anxiety, and general health status were evaluated. No significant differences regarding the BDI, BAI, GHQ-anxiety/insomnia, GHQ-social dysfunction, GHQ-severe depression, and total GHQ score were evident between cancer and control groups (P-value > 0.05). However, the Mann-Whitney test revealed a significant difference regarding GHQ-somatic symptoms score between chemotherapy-received patients and healthy controls (P-value = 0.019). The mean scores of GHQ-somatic symptoms were 23.71, 32.25, and 33.45 for control, AC-T, and TAC groups respectively. Comparison of GHQ-somatic symptoms scores between AC-T and TAC groups had shown no significant statistical difference (P-value = 0.714).

### 3.4 ACE-P

ACE-P was used in our study as a screening tool for cognitive impairment. Concerning this issue, the Kruskal-Wallis test was used which had shown no significant differences in ACE-memory, ACE-language, ACE-attention and orientation, ACE-fluency, ACE-visuospatial, MMSE, and ACE-total score among the mentioned three groups (AC-T, TAC, control). Further analyses were performed which revealed a significant difference between mean scores of the ACE-language of the AC-T and TAC groups (P-value = 0.036); in such a way that AC-T treated patients had worse performance in this test in comparison with the TAC group. Table 2 illustrates the summary of the ACE-P scores.

**Table 2:**
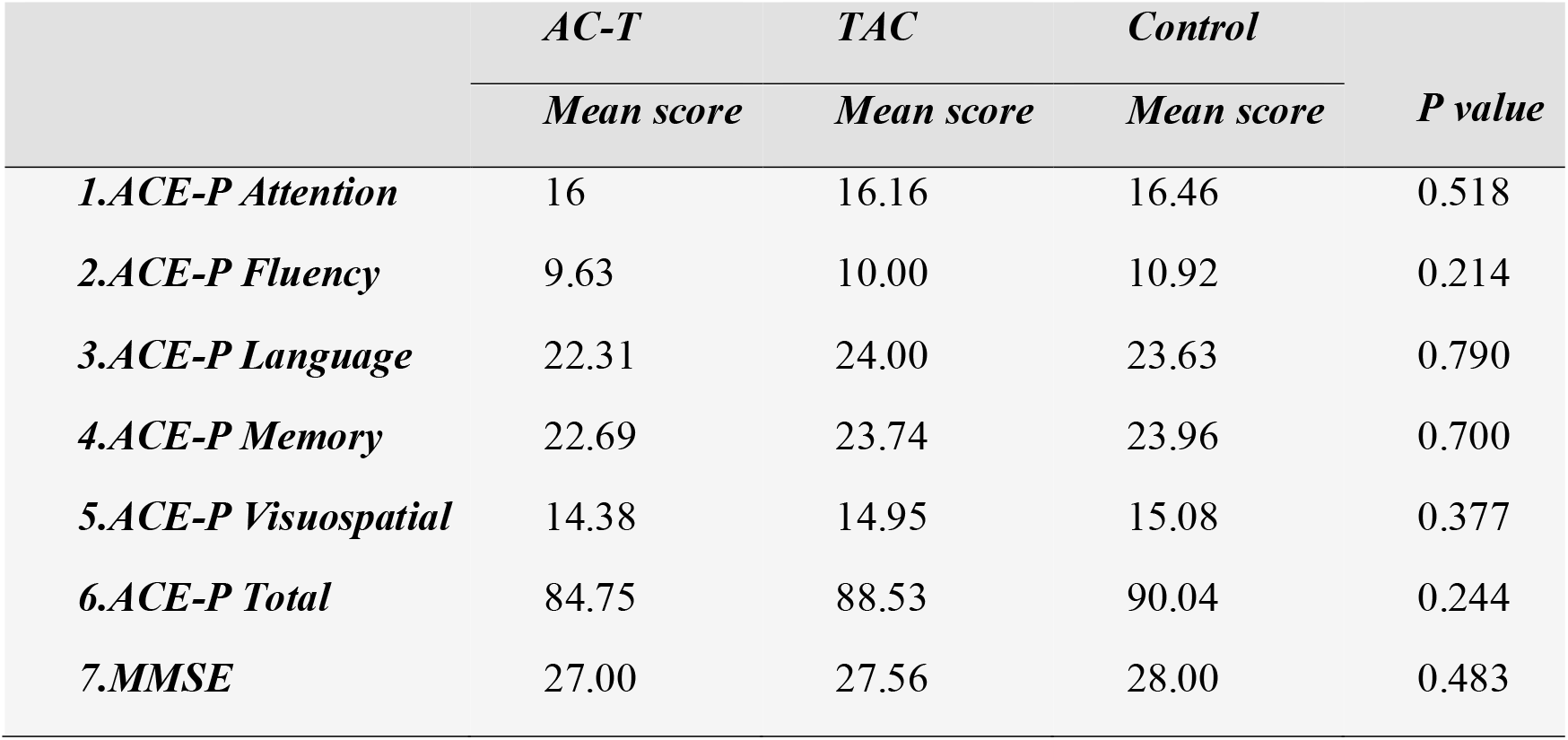
summary of mean scores of ACE-P scores.

|  | *AC-T* | *TAC* | *Control* |  |
| --- | --- | --- | --- | --- |
|  | *Mean score* | *Mean score* | *Mean score* | *P value* |
| **1.ACE-P Attention** | 16 | 16.16 | 16.46 | 0.518 |
| **2.ACE-P Fluency** | 9.63 | 10.00 | 10.92 | 0.214 |
| **3.ACE-P Language** | 22.31 | 24.00 | 23.63 | 0.790 |
| **4.ACE-P Memory** | 22.69 | 23.74 | 23.96 | 0.700 |
| **5.ACE-P Visuospatial** | 14.38 | 14.95 | 15.08 | 0.377 |
| **6.ACE-P Total** | 84.75 | 88.53 | 90.04 | 0.244 |
| **7.MMSE** | 27.00 | 27.56 | 28.00 | 0.483 |

### 3.5 CBS Battery

The CBS battery was our next tool for evaluating the participants’ cognition in aspects of memory, concentration, verbal ability, and reasoning. As the table 3 illustrated, among the CBS tasks performed in our study, no significant differences were shown regarding tasks of odd one out, digit span, rotation, polygons, feature match, paired associates, and spatial span between the three groups. However, the mean score of the double trouble test showed significant differences among the three groups. The AC-T treated patients got a significantly lower score in the double trouble task compared with the two other groups (P-value = 0.008). Besides, the AC-T regimen group had also shown significantly worse performance compared to the TAC regimen in the monkey ladder task (P-value = 0.031) through Mann-Whitney test but Kruskal-Wallis test did not show a significant difference among the three groups (P-value = 0.053). Finally, to create a metric representative of the overall cognitive function of each participant, Z-scores of the ten performed tasks were added together for each responder and then the mean Z-scores were compared between the three groups. It was demonstrated that the AC-T group overall had worse cognitive function compared with the two other groups (P-value=0.026). Although, the overall cognitive function did not show significant differences among the TAC and control groups (P-value=0.708). Table 3 and figure 1 present the summary of CBS battery scores.

**Table 3:** summary of CBS Battery scores. All scores have been converted to Z score values based on default CBS scores of normal populations.

| *CBS task* | *AC-T* | *TAC* | *Control* | *P value* |
| --- | --- | --- | --- | --- |
|  | *Mean*<br>*Z score* | *Mean*<br>*Z score* | *Mean*<br>*Z score* |  |
| *1.Digit span* | -1.419 | -1.183 | -0.967 | 0.312 |
| *2.Feature Match* | -1.486 | -1.379 | -1.388 | 0.919 |
| *3.Odd Ones Out* | -0.921 | -0.156 | 0.121 | 0.641 |
| *4.Rotation* | -1.150 | -0.821 | -0.644 | 0.162 |
| *5.Double Trouble* | -1.479 | -0.717 | -0.345 | 0.008 |
| *6.Monkey Ladder* | -1.675 | -1.101 | -1.350 | 0.053 |
| *7.Paired Association* | -1.519 | -1.412 | -1.280 | 0.740 |
| *8.Polygons* | -1.331 | -0.994 | -0.939 | 0.194 |
| *9.Spatial Span* | -1.600 | -1.173 | -1.386 | 0.156 |
| *10.Token Search* | -1.126 | -0.986 | -0.923 | 0.593 |
| *11.Overall cognitive status* | -1.264 | -0.871 | -0.809 | 0.026 |

**Figure 1:**
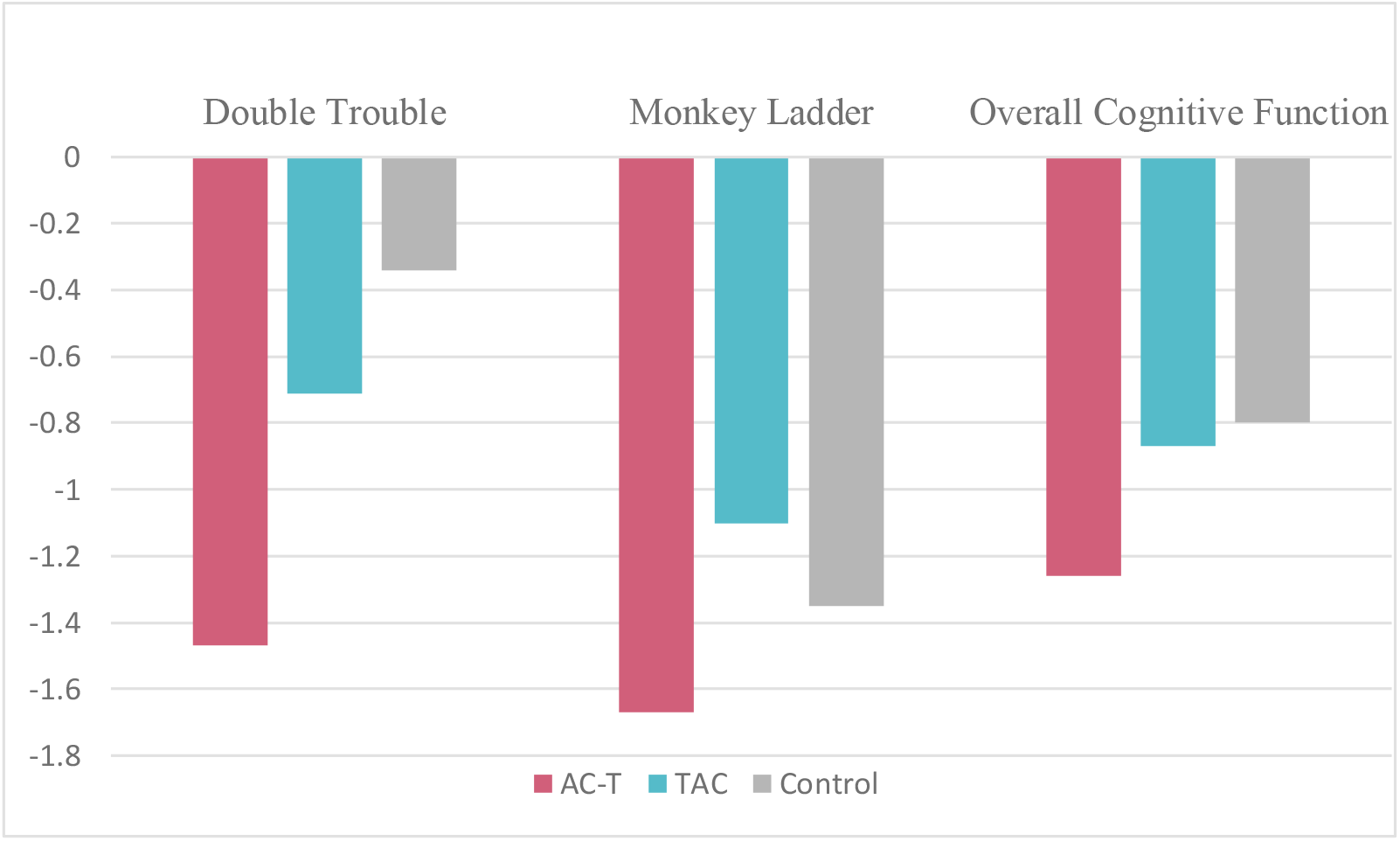
Mean Z-scores of double trouble test, monkey ladder test, and overall cognitive function regarding the CBS battery with significant differences among the three groups.

## 4. Discussion

As it is well-established, the phenomenon of “chemobrain” can affect BC survivors’ QoL. Therefore, it is important to select a less toxic chemotherapy regimen. In the current study, we aimed to identify any suspected cognitive impairment, depression, and anxiety in BC patients who received different chemotherapy regimens in order to compare their effects on patients’ cognitive function. Our study has revealed no significant differences between subjective symptoms of cognitive impairment, anxiety, and depression between BC patients and healthy individuals except for subjective fatigue which was higher in BC patients. In addition, Cognitive assessment regarding ACE-P tests has demonstrated poorer performance of the AC-T regimen group in comparison with the TAC regimen group in the language domain of cognition. Besides, the CBS battery results revealed worse cognitive status of AC-T treated patients associated with significant impairments in monkey ladder and double trouble tasks comapred with the two other groups.

As mentioned above, BC survivors reported more fatigue in comparison with the matched control group. Fatigue is one of the commonly reported complaints of cancer patients and is defined by National Comprehensive Cancer Network (NCCN) as a persistent sense of physical, emotional, or cognitive tiredness that interferes with patients’ usual function and is caused by cancer or its treatments [31]. Similar to our results, in a meta-analysis conducted by Abrahams et al., the pooled prevalence of severe fatigue was 27% among BC survivors which was significantly associated with higher stages of cancer and chemotherapy treatment; although this prevalence seemed to decrease about six months after treatment completion [32]. Besides, there are numerous studies that have demonstrated the association of cognitive impairment with fatigue [33, 34].

Furthermore, subjective cognitive impairments were similar in the three groups in the current study. This is in concordance with Pullens et al.’ study which showed inconclusive data regarding the higher prevalence of subjective cognitive impairment among BC patients in comparison with the normal population. They stated that this cognitive impairment cannot be related to cancer itself, chemotherapy, or hormonal therapy [35].

Analyzing BDI and BAI questionnaires showed no significant differences regarding depression and anxiety between BC patients and the control group. A study conducted by Hadi et al. confirmed our results. They recruited 178 BC patients at least one year after the cancer diagnosis and 400 healthy women. The two groups did not show significant differences regarding depression and anxiety [36]. However, some studies revealed different results. In a study conducted by Shilling et al., about 76 % of BC patients reported anxiety six months after chemotherapy compared with 57% of BC patients who did not receive any chemotherapy regimens. Also, patients with higher anxiety rates had worse performance in memory-related tasks [37]. Cheung et al. found similar results. Chemotherapy-treated BC patients had higher prevalence of anxiety compared to non-chemotherapy patients (21.9% vs 8.6%; p-value = 0.002) [38]. Depression was also among the significantly affected domains in some experiments. Ibrahim et al. conducted a meta-analysis that revealed increased depression rates in patients receiving taxane-based chemotherapy compared to other regimens and healthy controls [39]. Zunini et al. in 2012 also used BDI and BAI, same as our study, to estimate mood disorders in BC patients. They found that chemotherapy-treated patients significantly differ from the control group in BDI and BAI scores [40].

Taken together, results regarding the prevalence of mood disorders in cancer patients are inconclusive. Besides, cognitive impairment observed in BC patients can be related to the associated psycho-social disorders including stress, depression, and anxiety besides neurotoxic sequela of the underwent treatments. Finding no significant differences in terms of depression and anxiety between the three groups participating in our study would make the cognitive assessments more reliable. Identifying any significant cognitive impairments among the three groups can be attributed to differences in underwent treatments and in particular chemotherapy agents after removing the confounding effects of concurrent psychological disorders.

Despite the BDI and BAI, GHQ-somatic symptom scores were significantly higher in BC patients. Similarly, several studies have confirmed a high prevalence of somatization or somatic symptoms including sleep disturbance, pain, changes in libido and appetite, fatigue, etc. in cancer patients affecting their QoL. In a study conducted by Palesh et al. in 2010, up to 80% of cancer patients reported sleep disturbance after the first cycle of chemotherapy compared to the 30% in the general population. Among them, BC patients had the highest number of sleep disturbance and insomnia complaints [41]. A systematic review by Andersen et al. in 2011 showed chronic pain syndrome in 25 to 60% of BC patients after treatment [42]. Another study by Coates et al. recruited 100 cancer patients to score the severity of some post-chemotherapy complaints. Feeling sick was the top complaint with the highest prevalence and severity score among cancer patients [43].

However, these somatic complaints can be resulted from cancer itself, the treatment, or concurrent psychological disorders such as depression. As Caruso et al. stated, somatic complaints can cause increased cancer-related disability, poor compliance with treatment protocols, delay in recovery, poor outcome, and reduced QoL. Besides, the overlap of somatic symptoms caused by consequences of cancer, received treatments such as chemotherapy, and depressive/anxiety disorders can challenge identifying the exact underlying etiology [44]. Therefore, since our results did not reveal significant differences between AC-T, TAC, and control groups in terms of anxiety and depression, this higher magnitude of somatic symptoms in cancer and in particular TAC group can be related to consequences of the underwent chemotherapy treatments besides underlying cancer.

Using the ACE-P inventory for assessment of cognitive function has revealed no significant differences between BC patients and healthy individuals. However, the AC-T group had poorer cognitive functioning in the language domain in comparison with the TAC group. Various studies have demonstrated language as one of the cognitive domains which are impaired in BC patients relative to healthy controls [45, 46]. In a meta-analysis conducted by Bernstein et al., language, attention, processing speed, immediate recall, delayed recall, and executive function were shown to be impaired in chemotherapy-received patients compared to healthy control. Although, after adjustment for pre-chemotherapy cognitive differences, only memory recall and executive function remained altered. It is noteworthy that this research did not reveal any chemotherapy effect on cognition after comparison of survivors who had received chemotherapy and those who had not [47]. Despite this research, we did not find significant differences between BC patients and healthy controls in the language domain; however, the AC-T group showed significantly worse performance compared with the TAC-treated patients in this domain suggesting the possible toxic effect of this regimen on verbal function.

In addition, CBS was used for a detailed evaluation of different aspects of cognitive domains which revealed the worse performance of patients treated with the AC-T regimen in tasks of double trouble and monkey ladder compared with the other groups.

Double trouble task measures sustained focused attention and response inhibition, a subset of concentration which means focusing on relevant data to make an appropriate response despite distracting stimuli [30]. Sustained attention depends on activation of task-relevant processes and inhibition of task-irrelevant processes (response inhibition). These aspects of concentration are mainly regulated through two distinct neural networks, dorsal and ventral attention networks. Dorsal attention network (DAN), mediates top-down voluntary allocation of attention and ventral attention network (VAN) regulates detection of irrelevant stimuli and shifting attention or response inhibition [48]. However, some studies have also highlighted the role of DAN in the feature-based suppression of task-irrelevant information besides the VAN [49]. Monkey ladder test requires creating a set of relationships between some objects in the space which involves visuospatial working memory. As mentioned in a study conducted by Majerus et al., DAN is also involved in both verbal and visual working memory through controlling both quantitative and qualitative aspects of attention during working memory tasks [50].

Therefore, results revealed the worse performance of AC-T-treated BC patients in the attention domain in comparison with TAC-treated cases and healthy volunteers. Similarly, Jansen et al. recruited BC patients planned to receive the AC-T regimen before, during, and after chemotherapy. They found the decreased performance of the participants in the attention domain during chemotherapy; however, the impairment was improved significantly in long-term follow-ups [16]. Also, in another study conducted by Chen et al., chemotherapy-received BC patients had lower scores in some cognitive domains including attention via Attention Network Test [51]. However, Jenkins et al. found inconsistent results. They showed that few BC women had measurable attention deficits while the majority were unaffected [9]. Despite studies evaluating attention deficits in BC patients, scant articles have assessed the magnitude of impairment in different regimens to propose the most toxic agents. Therefore, our results highlighted potential toxic sequela of AC-T regimen on dorsal and ventral attention networks reflected as impairments in attention-related tasks. Performing further neuroimaging studies to confirm functional or structural cortical changes in DAN and VAN neural networks in AC-T-treated patients would be the next research priority.

In conclusion, although this study has confirmed the chemobrain concept similar to previous literature and revealed potential worse neurotoxic effects of the AC-T regimen on attention, working memory, and language domains compared with the other commonly prescribed regimen, the exact mechanism behind the chemobrain has not been fully identified and further research is needed. Since the emergence of the chemobrain concept, different mechanisms have been proposed for this phenomenon. Neurotoxicity is a well - known side effect of taxanes in such a way that they can cause peripheral neuropathy, ataxia, cognitive decline, etc. Paclitaxel and docetaxel, which are parts of the AC-T and TAC regimens respectively, are the most commonly used taxanes. The mechanisms suggested for their neurotoxic impacts despite limited crossing of the blood-brain barrier (BBB) include negative effects on microtubule dynamics and synaptic plasticity, neuronal apoptosis, decreased hippocampal cell proliferation, cytokine dysregulation, and consequently neuroinflammation [52]. In addition, as most of the BC chemotherapeutics with the exception of cyclophosphamide and fluorouracil can’t easily cross the BBB, the oxidative stress and cytokine dysregulation seem to play an important role in developing chemobrain [53, 54].

Nevertheless, our study has some limitations. Due to high psychosocial stress in patients who have just been diagnosed with BC, recruitment of the patients for assessment of the baseline cognitive parameters before receiving the chemotherapy was not feasible in our settings. Moreover, as part of data gathering coincided with the global COVID-19 pandemic, we were not able to increase the number of patients considering their health issues. Although we have tried to consider psychosocial problems, further longitudinal research is needed to reinforce our findings, especially by neuroimaging studies such as fMRI.

## 5. Conclusion

In conclusion, although the BC patients seem to be not aware of their cognitive impairment, the AC-T receiving survivors had poorer performance in tasks involving visuospatial working memory and concentration. To limit the confounding effects of psychosocial problems on cognitive function, depression and anxiety were assessed which revealed no significant differences between the BC survivors and healthy controls. However, BC patients complained of chronic fatigue which may affect cognition, independently. Yet, the two mentioned chemotherapy regimens did not differ in chronic fatigue. Overall, the current study suggests prescribing the AC-T regimen with caution in patients suffering from baseline cognitive impairment and recommends choosing the TAC regimen as an alternative.

## Data Availability

All data generated or analyzed during this study are available from the corresponding author on reasonable request.

## Acknowledgements

The authors would like to thank DANA Brain Health Institute; Iranian Neuroscience Society, Fars Chapter, Shiraz, Iran for the received support.

## Declarations

### Funding

Partial financial support was received from School of Advanced Medical Sciences and Technologies, Shiraz University of Medical Sciences, Shiraz, Iran (ID: 15307)

### Conflicts of interest/Competing interests

The authors have no conflicts of interest to declare that are relevant to the content of this article.

### Availability of data and material

All data generated or analysed during this study are available from the corresponding author on reasonable request.

### Code availability

Not applicable.

### Authors’ contributions

All authors contributed to the study conception, design, and read and approved the final manuscript.

## Compliance with Ethical Standards

### Ethics approval

All procedures performed in studies involving human participants were in accordance with the ethical standards of the institutional and/or national research committee and with the 1964 Helsinki Declaration and its later amendments or comparable ethical standards. The study was approved by the Ethics Committee of the Shiraz University of Medical Sciences, Shiraz, Iran.

### Consent to participate

Informed consent was obtained from all individual participants included in the study.

### Consent for publication

Additional informed consent was obtained from all individual participants for whom identifying information is included in this article.

## Notes

### Competing Interest Statement

The authors have declared no competing interest.

### Funding Statement

Partial financial support was received from the School of Advanced Medical Sciences and Technologies, Shiraz University of Medical Sciences, Shiraz, Iran (ID: 15307)

